# Knowledge, attitudes and practices regarding antibiotic use and antimicrobial resistance among community pharmacy workers in Bukavu, Eastern Democratic Republic of Congo: a cross-sectional study

**DOI:** 10.64898/2026.11.16.26368184

**Authors:** Willy Mushobekwa Biringanine, Nancy Rugajo, Justin Ntibonera Aganze, Octave Cito Mudekereza, Corneille Kabagale Ansima, Lameck Zigashane Kawala, Emmanuel Lurhangire Ongezi, David Lupande Mwenebitu

## Abstract

**Background:** Community pharmacies are often the first point of care in low- and middle-income countries, yet the knowledge, attitudes and practices (KAP) of their workers regarding antibiotics and antimicrobial resistance (AMR) are undocumented in the Democratic Republic of Congo (DRC).

**Objectives:** To assess KAP regarding antibiotic use and AMR, and their determinants, among community pharmacy workers in Bukavu, Eastern DRC.

**Methods:** A cross-sectional survey was conducted in April–June 2026 among 423 community pharmacy workers, one per pharmacy, selected by multistage sampling across the three health zones of Bukavu, using a structured, interviewer-administered questionnaire. Knowledge (8 items), attitudes (9 items) and dispensing practices (4 items) were scored using modified Bloom’s cut-offs. Multivariable logistic regression identified determinants.

**Results:** Good knowledge was found in 19.4%, favourable attitudes in 17.5% and appropriate dispensing practices in 27.4%. Overall, 96.5% dispensed antibiotics without a prescription and 9.2% knew the WHO AWaRe classification. University education and pharmaceutical qualification were independently associated with good knowledge [adjusted OR (aOR) 4.38 and 2.41] and appropriate practices (aOR 5.74 and 2.01). Good knowledge was associated with favourable attitudes (aOR 4.86, 95% CI 2.68–8.82), which in turn predicted appropriate practices (aOR 6.62, 95% CI 3.50–12.53). In addition, 57.0% knew of “Machange”, an informal mixture of medicines: 99.6% of those describing its composition listed at least one antibiotic, and 65.6% of those familiar with it sold it.

**Conclusions:** Community pharmacy workers in Bukavu show substantial gaps in antibiotic-related KAP. Stewardship interventions should combine education with regulation, diagnostic capacity and AMR surveillance within a One Health framework.

## Introduction

Antimicrobial resistance (AMR) is one of the defining public health challenges of the 21st century, threatening decades of progress in the prevention and treatment of infectious diseases and undermining the effectiveness of life-saving antimicrobial therapies.^1,2^ In 2021, bacterial AMR was directly responsible for an estimated 1.14 million deaths worldwide and contributed to nearly 4.7 million deaths overall, with the mortality burden concentrated in low- and middle-income countries (LMICs), where infectious diseases remain common.^1^ In Africa, systematic reviews have demonstrated widespread resistance to commonly used antibiotics across both hospital and community settings.^3^ Beyond its human cost, AMR places a growing burden on health systems by prolonging hospital stays, escalating healthcare expenditure, and narrowing therapeutic options, thereby jeopardizing progress toward universal health coverage and sustainable development.^2,4^

Antibiotic overuse, including inappropriate prescribing, self-medication, improper dispensing, and poor treatment adherence, remains one of the main preventable drivers of AMR.^1,2,5^ Optimizing antibiotic use at every level of healthcare is therefore a cornerstone of global strategies to preserve the effectiveness of existing antibiotics and limit the future consequences of resistance.^6,7^ The One Health approach, which recognizes the interconnectedness of human, animal and environmental health, is increasingly promoted as an essential framework for combating AMR, as resistant organisms and resistance genes circulate across these domains.^2,6,7^ However, awareness of the One Health concept remains limited among many medical professionals, and increasing its dissemination is crucial for effective AMR control and prevention.^8^ As recently emphasized, the One Health approach is particularly important for addressing neglected zoonotic diseases, which are linked to antimicrobial resistance and require coordinated strategies across sectors.^9^

Community pharmacies are key players in health systems worldwide and are often the first point of contact for people seeking treatment, particularly in LMICs where access to formal healthcare is limited.^10–13^ Positioned between health systems and communities, pharmacy workers are well placed to promote rational antibiotic use through dispensing, counselling, and referral,^11,12^ making them increasingly recognized as partners in antimicrobial stewardship (AMS).^11,13^ However, these roles depend on pharmacy workers having adequate knowledge, favourable attitudes, and sound dispensing practices regarding antibiotics and AMR.^7,14^

LMICs bear a disproportionate share of the AMR burden due to a high prevalence of infectious diseases combined with weak healthcare delivery and pharmaceutical regulation systems.^1,2^ Limited access to affordable medical consultations and shortages of qualified health professionals have made community and informal pharmacies one of the most accessible sources of antibiotics in these settings.^11,15–17^ High out-of-pocket healthcare costs often lead individuals to bypass formal consultations and obtain antibiotics directly from pharmacies without prior clinical evaluation, increasing the risk of inappropriate use.^1,2,13,18^

At the same time, AMS in LMICs remains constrained by limited laboratory capacity, weak AMR surveillance, poor regulatory enforcement, and insufficient financial and human resources.^2,7^ These challenges highlight the need for robust evidence on antibiotic-related knowledge, attitudes, and practices among community pharmacy workers, particularly in settings where pharmacies play a central role in healthcare access.^11,13^

The Democratic Republic of Congo (DRC) exemplifies many of these structural challenges. Despite a heavy burden of infectious diseases, the country faces a persistent shortage of qualified health professionals, limited diagnostic capacity, weak health system governance, and recurrent economic crises, population displacement, and armed conflict, all of which strain an already fragile healthcare system.^2,3,19^ Healthcare financing continues to rely heavily on out-of-pocket payments, making consultations unaffordable for much of the population and driving many to seek treatment directly from community pharmacies.^20–22^ In Bukavu, community pharmacies have proliferated rapidly in response to this demand, yet regulatory oversight remains weak, and access to diagnostic and antimicrobial susceptibility testing is limited. Recent microbiological data from Bukavu have documented high resistance rates among clinically important pathogens, confirming that irrational antibiotic use is a documented, not theoretical, problem.^23,24^ Consequently, pharmacies have become one of the most accessible entry points into the healthcare system, particularly for managing common infectious conditions, extending well beyond their traditional dispensing role.^1,3,11,12^ Yet the rapid growth of community pharmacies, combined with weak enforcement of pharmaceutical regulation and AMS, raises concerns about the quality of antibiotic dispensing and its contribution to inappropriate antibiotic use and AMR.^2,13,14^

While numerous studies have assessed healthcare professionals’ knowledge, attitudes, and practices regarding antibiotic use and AMR, evidence remains uneven across regions, with most research concentrated in Asia, the Middle East, and a limited number of African countries.^11,13,17,25,26^ Data from Central Africa are particularly scarce, and few studies have specifically examined community pharmacy workers despite their growing role in antibiotic access and dispensing.^11,15^ In the DRC, existing research has largely focused on hospital, prescriber, or population-level settings, leaving community pharmacy workers’ knowledge, attitudes, and practices (KAP), especially in the eastern part of the country, poorly documented.^19,23,24,27^ Previous studies in the DRC have reported gaps in healthcare providers’ knowledge of antibiotic use and resistance in hospital settings in Kinshasa,^27^ while population-level and prescriber-focused studies have documented widespread inappropriate antibiotic use. However, no study to date has comprehensively assessed KAP among community pharmacy workers in Eastern DRC, and the specific pathways through which these workers influence antibiotic access—including dispensing without prescriptions, patient counselling, and referral practices—remain poorly understood. This evidence gap limits the ability to design context-specific antimicrobial stewardship interventions and strengthen the role of community pharmacies in preventing AMR.^2,12^

Against this background, this study assessed KAP toward antibiotic use and AMR among community pharmacy workers in Bukavu, Eastern DRC, and explored the determinants of good knowledge, favourable attitudes, and appropriate practices. By generating context-specific evidence in a setting where community pharmacies are integral to primary healthcare access, this study aims to address a significant gap in the literature and inform AMS initiatives, strengthen pharmaceutical practice, and support evidence-based public health policy in the DRC and other resource-limited settings.

## Methods

### Ethics

The study protocol was reviewed and approved by the Ethics Committee of the Official University of Bukavu (UOB) (approval number UOB/CEM/016/2026). The study was conducted in accordance with the Declaration of Helsinki^28^ and with national and institutional standards. In the DRC, any university institution authorized by the government may grant ethical approval independently of the authors’ affiliations. This study is part of a larger One Health research programme aimed at improving knowledge of AMR and characterizing Gram-negative bacilli resistant to third-generation cephalosporins, carbapenems and colistin. Data collection was undertaken only with the ethical and administrative permission of the Central Officers of the three health zones of Bukavu, and approval was also obtained from the managers of all selected community pharmacies. Participation in the study was entirely voluntary. Before each interview, participants were informed about the study’s purpose and procedures, and written informed consent was obtained. Participants were informed of their right to decline participation or withdraw from the study at any time without penalty. To ensure anonymity, no personal identifiers were collected, and all questionnaires were anonymized prior to data entry and analysis. Data were stored securely and were accessible only to the research team.

### Study design

The study was a cross-sectional survey conducted among community pharmacy workers in Bukavu, Eastern DRC, between April and June 2026. It aimed to assess participants’ KAP regarding antibiotic use and AMR, and to identify factors associated with good knowledge, favourable attitudes and appropriate practices. The study was designed and reported in accordance with the Strengthening the Reporting of Observational Studies in Epidemiology (STROBE) guidelines for cross-sectional studies (completed checklist in Supplementary File S1).

### Study setting

The study was carried out in Bukavu, the capital of South Kivu Province in eastern DRC. Bukavu is divided into three health zones (Ibanda, Kadutu and Bagira), each comprising several health areas that together form the administrative framework for healthcare delivery. The geographic distribution of the surveyed community pharmacies across the three health zones is shown in Figure 1. As one of the largest urban centres in eastern DRC, Bukavu is characterized by a high burden of infectious diseases, rapid urbanization, and many community pharmacies that play a critical role in ensuring access to healthcare and medicines. Community pharmacies are frequently the first point of care sought for common illnesses, making Bukavu an appropriate setting for investigating knowledge, attitudes and practices related to antibiotic use and AMR among community pharmacy workers.

**Figure 1.** Geographic distribution of surveyed community pharmacies in the three health zones of Bukavu, Eastern DRC. Each marker represents one of the 423 surveyed community pharmacies; grey shading with black lines shows the boundaries of the health areas of the Ibanda, Kadutu and Bagira health zones. Inset maps show the location of Bukavu within South Kivu Province, the DRC and Africa.

### Study population

The study population comprised workers employed in community pharmacies across the three health zones (Ibanda, Kadutu and Bagira) of Bukavu. Eligible participants were those actively involved in dispensing medicines or providing pharmaceutical services to clients, regardless of professional qualification (pharmacists or non-pharmacist personnel).

### Eligibility criteria

Community pharmacy workers were eligible to participate if they were employed in a community pharmacy located in Bukavu during the study period, were involved in dispensing medicines or providing pharmaceutical services to clients, and voluntarily agreed to participate after providing written informed consent. Workers who were temporarily absent during the survey period, declined to participate, or were not directly involved in pharmaceutical service provision were excluded from the study.

### Sample size determination

The single population proportion formula (Cochran’s formula, also referred to as Schwartz’s formula) was used to compute the minimum sample size:

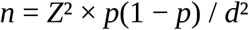

Where n denotes the required sample size, Z the standard normal deviate at a 95% confidence level (1.96), p the expected proportion of workers with good KAP (50%, chosen because no previous local data were available and because this value maximizes the sample size) and d the margin of error (5%). This yielded a minimum sample size of 384. To allow for an anticipated 10% non-response, the target was increased to 423 participants (384 × 1.10). Because one worker was interviewed per pharmacy, this target also corresponded to 423 community pharmacies. All 423 community pharmacy workers approached agreed to participate (response rate 100%) and all were included in the analysis. For the multivariable logistic regression models, the number of outcome events per predictor variable, among the 417 participants with complete data, was 13.5 for the knowledge model (81 events, 6 predictors), 10.6 for the attitudes model (74 events, 7 predictors) and 14.5 for the practices model (116 events, 8 predictors), all at or above the commonly recommended minimum of 10.

### Sampling procedure

Because no comprehensive and up-to-date registry of community pharmacies existed in Bukavu, a multistage sampling strategy stratified by health zone and health area was used. Stage 1 (health zones and health areas): all three health zones of Bukavu (Ibanda, Kadutu and Bagira) and all of their health areas (37 health areas in total) were included. Stage 2 (allocation): the target sample of 423 pharmacies was allocated to each health area in proportion to its population, using population figures from the 2025 South Kivu Provincial Health Report. An initial list of community pharmacies was obtained from the South Kivu Provincial Health Division; however, local health authorities acknowledged that this list was incomplete because of the rapid and largely uncontrolled expansion of community pharmacies across the city. It was therefore used for planning only and not as the sampling frame.

Stage 3 (selection of pharmacies): in each health area, field teams first listed all operating community pharmacies by systematically walking through the health area, thereby building an on-site sampling frame. The number of pharmacies allocated to that health area was then drawn from this list by simple random sampling without replacement, using a random number generator. A selected pharmacy that was closed on two visits or had no eligible worker present was replaced by the next pharmacy randomly drawn from the same list. Where fewer pharmacies were operating than had been allocated, all operating pharmacies in the health area were included and the shortfall was reallocated to the other health areas of the same health zone in proportion to their population. Stage 4 (selection of participants): one worker was interviewed per pharmacy; if more than one eligible worker was present, one was selected at random by lottery. Each of the 423 participants therefore came from a different pharmacy, and the number of pharmacies equals the number of participants. Two health areas (Ciguri and Mushekere) had no operating community pharmacy at the time of the survey and therefore contributed no participants; the remaining 35 health areas contributed between 3 and 40 pharmacies each. The final sample comprised 423 pharmacies (and 423 workers): Ibanda (n = 146; 34.5%), Kadutu (n = 182; 43.0%) and Bagira (n = 95; 22.5%). The planned and achieved sample sizes for each health area are given in Supplementary File S2, and the locations of the surveyed pharmacies are shown in Figure 1.

### Study variables

The main outcome variables were participants’ KAP concerning antibiotic use and AMR, assessed using eight, ten and seven items, respectively, of which eight, nine and four items were retained in the final scores after reliability analysis (see Assessment of internal consistency). Explanatory variables were age, sex, highest educational level, professional qualification, years of professional experience and prior training on antibiotic use and antimicrobial resistance. For analysis, educational level was dichotomized as primary/secondary (n = 112) or university or above (n = 311), and professional qualification as pharmaceutical (pharmacists, pharmacy technicians (G3) and pharmacy assistants (A2); n = 110) or non-pharmaceutical (nurses, physicians, laboratory technicians, midwives and other personnel; n = 313).

### Data collection instrument

Data were collected through a structured, interviewer-administered questionnaire developed from previously validated KAP questionnaires on antibiotic use and antimicrobial resistance and adapted to the Congolese context following a review of the relevant literature. The questionnaire included sections on socio-demographic and professional characteristics; KAP related to antibiotic use and AMR; and items on the dispensing and composition of locally known mixed antibiotic preparations (“Machange”). The questionnaire was administered in French. Prior to data collection, it was reviewed by public health experts and pre-tested among 10 community pharmacy workers who were not included in the study sample, for clarity, relevance and contextual appropriateness. The full questionnaire, with the wording of every item, its response options and the scoring key, is provided in Supplementary File S3.

### Data collection procedures

Data were collected between April and June 2026 by trained research assistants through face-to-face interviews in selected community pharmacies using KoboCollect tools. Study objectives were explained individually to each eligible participant, and written informed consent was obtained before each interview. Interviews were conducted in the most private area of the pharmacy practicable, to ensure confidentiality and minimize interruptions. All eligible participants completed the questionnaire.

### Assessment of KAP

Knowledge was assessed using eight items covering antibiotic use, indications, dispensing and AMR; attitudes using ten items on participants’ perceptions of, and professional attitudes towards, antibiotic dispensing and AMS; and antibiotic-dispensing practices using seven self-reported items on routine dispensing behaviour. All items are listed in Table 2. Each item was scored as 1 for a correct (knowledge), favourable (attitude) or appropriate (practice) response and as 0 otherwise; “don’t know” and “prefer not to answer” responses were scored 0. For expired antibiotics, only returning them to the health zone central office was considered appropriate, and for hand hygiene only the answer “yes” (as opposed to “not very often” or “no”). Domain scores were the sum of the retained items only (eight knowledge, nine attitude and four practice items; see below). Participants were classified as having good knowledge (≥80% of the maximum score, ie ≥7 of 8 items), favourable attitudes (≥60%, ie ≥6 of 9 items) or appropriate practices (≥75%, ie ≥3 of 4 items); all others were classified as having poor knowledge, unfavourable attitudes or inappropriate practices, respectively. A lower threshold was used for attitudes because the highest observed attitude score was 8 of 9 and only 10 participants (2.4%) reached the 80% level, which would not have allowed multivariable analysis. Non-prescription dispensing was explored with two distinct questions: whether the participant dispensed antibiotics without a prescription (and, if so, how often), and whether there were specific antibiotics that the participant never dispensed without a prescription (and, if so, which). These questions measure different things: a participant could restrict a few antibiotics to prescription-only dispensing while dispensing others without a prescription.

### Assessment of internal consistency

The internal consistency of the knowledge, attitude and practice scales was assessed using Cronbach’s alpha. Items with a corrected item–total correlation below 0.10 were excluded from the final scores. All eight knowledge items met this criterion and were retained (α = 0.707). For attitudes, one item (not stopping antibiotic treatment once symptoms improve; item–total correlation 0.05) was excluded, increasing alpha from 0.692 to 0.710 and leaving nine items. For practices, three items (not dispensing antibiotics without a prescription, not substituting a prescribed antibiotic, and adapting dispensing to children and adults; item–total correlations 0.06–0.09) were excluded, increasing alpha from 0.521 to 0.652 and leaving four items: checking expiry dates, never dispensing at least one antibiotic without a prescription, returning expired antibiotics to the health zone central office, and practising hand hygiene during working hours. Final domain scores were calculated using the retained items only. Excluded items are still reported descriptively in Table 2, where they are marked with a dagger (†); item-level statistics are given in Supplementary File S4.

### Statistical analysis

Data were entered, cleaned and analysed using Stata/MP version 17.0 (Stata Corp LLC, College Station, TX, USA). Categorical variables were summarized using frequencies and percentages, and continuous variables were described using means and standard deviations or medians and interquartile ranges (IQRs). Normality of continuous variables was examined using the Shapiro-Wilk test and graphical methods. For categorical variables, comparisons between groups were performed using Pearson’s chi-square test or Fisher’s exact test; for non-normally distributed continuous variables, the Wilcoxon rank-sum test was used. Bivariate associations between each candidate variable and each outcome are reported in Supplementary File S5. Multivariable logistic regression was used to identify independent determinants of good knowledge, favourable attitudes and appropriate practices. Covariates were specified a priori on the basis of the literature (age, sex, educational level, years of professional experience, professional qualification and prior training) and the same set was kept in all three models, irrespective of bivariate *P* values, to allow comparison across domains; knowledge level was added to the attitudes model, and knowledge and attitude levels to the practices model. Individual KAP items were not entered as predictors, because they are the components from which the outcome scores were derived. Health zone was not included in the main models; a sensitivity analysis adding it is reported in Supplementary File S5. Analyses were complete-case: age was missing for 6 participants (1.4%), so the multivariable models included 417 participants; no other variable had missing values.

The final models included the following covariates (Table 3):

**Knowledge model:** age, sex, educational level, years of professional experience, professional qualification and prior training on antibiotic use and AMR (6 predictors; 81 events).

**Attitudes model:** the six covariates of the knowledge model plus knowledge level (good vs poor) (7 predictors; 74 events).

**Practices model:** the six covariates of the knowledge model plus knowledge level and attitude level (favourable vs unfavourable) (8 predictors; 116 events).

Multicollinearity was assessed using variance inflation factors (mean 1.38 for knowledge, 1.35 for attitudes and 1.35 for practices; all within acceptable limits). Model fit was assessed using the Hosmer-Lemeshow test: knowledge χZ = 4.28, *P* = 0.831; attitudes χZ = 5.37, *P* = 0.717; practices χZ = 9.87, *P* = 0.275; all indicating good fit. Continuous variables (age and professional experience) were retained as continuous variables. Reference categories were: female for sex, primary/secondary for education, no for training, non-pharmaceutical for profession, poor knowledge for knowledge level, and unfavourable attitude for attitude level. Adjusted odds ratios (aORs) with 95% confidence intervals (95% CI) were reported, and statistical significance was set at *P* < 0.05. All regression models consistently reported aORs, 95% CIs and *P* values.

## Results

### Sociodemographic and professional characteristics of community pharmacy workers

Table 1 summarizes the socio-demographic and professional characteristics of the 423 community pharmacy workers who participated in this study. The median age was 28 years (IQR 24–34; age missing for 6 participants), and the median duration of professional experience was 3 years (IQR 1–5). Most participants were women (65.0%). Most participants had a university-level or postgraduate education (n = 311; 73.5%), whereas 112 (26.5%) had primary or secondary education. Nurses were the largest professional group (51.8%), followed by other personnel (15.1%), pharmacy technicians (13.5%) and pharmacists (7.1%); overall, 110 participants (26.0%) held a pharmaceutical qualification. Participants were recruited from the three health zones of Bukavu, with the largest proportion from Kadutu (43.0%), followed by Ibanda (34.5%) and Bagira (22.5%). Overall, 66.4% of participants had previously received training in antibiotic use and antimicrobial resistance.

**Table 1.** Sociodemographic and professional characteristics of community pharmacy workers in Bukavu, Eastern DRC (*N* = 423)

| **Characteristic** | **n (%)** | **Median (IQR)** |
|---|---|---|
| Age (years) |  | 28 (24–34) |
| Sex |  |  |
| Female | 275 (65.0) |  |
| Male | 148 (35.0) |  |
| Professional experience (years) |  | 3 (1–5) |
| Highest educational level |  |  |
| Primary | 2 (0.5) |  |
| Secondary | 110 (26.0) |  |
| University | 309 (73.0) |  |
| Postgraduate | 2 (0.5) |  |
| Professional qualification |  |  |
| Pharmacist | 30 (7.1) |  |
| Pharmacy technician (G3) | 57 (13.5) |  |
| Pharmacy assistant (A2) | 23 (5.4) |  |
| Physician | 17 (4.0) |  |
| Nurse | 219 (51.8) |  |
| Laboratory technician | 4 (1.0) |  |
| Midwife | 9 (2.1) |  |
| Other<sup>a</sup> | 64 (15.1) |  |
| Health zone |  |  |
| Bagira | 95 (22.5) |  |
| Ibanda | 146 (34.5) |  |
| Kadutu | 182 (43.0) |  |
| Previous training on antibiotic use and AMR |  |  |
| Yes | 281 (66.4) |  |
| No | 142 (33.6) |  |

IQR, interquartile range (25th–75th percentile); AMR, antimicrobial resistance. Age was missing for 6 participants (median calculated on n = 417).
<sup>a</sup>Secondary-school pupils (n = 31), students (8), public health graduates (7), no profession (6), housewives (3), economists (2), agronomists (2), first-aid workers (2), and one each of lawyer, nutritionist and geologist.

In summary, the study population comprised predominantly young, female community pharmacy workers with relatively limited professional experience, high levels of formal education, and a relatively high proportion with prior training in antibiotic use and antimicrobial resistance.

### KAP regarding antibiotic use and AMR among community pharmacy workers

The participants’ knowledge, attitudes and practices are summarized in Table 2. Overall classifications are based on the retained items only; items excluded after reliability analysis are marked †.

**Table 2.** KAP regarding antibiotic use and AMR among community pharmacy workers in Bukavu, Eastern DRC (*N* = 423)

| **Domain/item** | **Favourable response, n (%)** |
|---|---|
| **Knowledge** |  |
| Awareness of the WHO AWaRe classification | 39 (9.2) |
| Recognized excessive antibiotic use as a driver of AMR | 349 (82.5) |
| Recognized that AMR can make infections difficult to treat | 338 (79.9) |
| Knowledge of antimicrobial susceptibility testing and its purpose | 155 (36.6) |
| Recognized self-medication as a driver of AMR | 344 (81.3) |
| Recognized hand hygiene as an effective measure to prevent infections | 315 (74.5) |
| Recognized that inappropriate antibiotic use may worsen disease outcomes | 389 (92.0) |
| Knowledge of the pharmacological classification of antibiotics | 171 (40.4) |
| **Overall: good knowledge** | **82 (19.4)** |
| **Overall: poor knowledge** | **341 (80.6)** |
| **Attitudes** |  |
| Did not routinely recommend antibiotics to clients | 7 (1.7) |
| Agreed that antibiotics should only be dispensed with a prescription | 171 (40.4) |
| Disagreed that personal experience alone is sufficient to dispense antibiotics | 91 (21.5) |
| Did not support dispensing antibiotics for fever, cold or cough lasting <2 days | 164 (38.8) |
| Did not support dispensing antibiotics for acute diarrhoea lasting <1 day | 121 (28.6) |
| Did not support dispensing antibiotics for cough alone | 94 (22.2) |
| Recognized that influenza does not routinely require antibiotics | 132 (31.2) |
| Recommended medical consultation before antibiotic use | 190 (44.9) |
| Considered regular regulatory inspections of community pharmacies important | 362 (85.6) |
| Agreed that antibiotic treatment should not be discontinued once symptoms improve<sup>a</sup> | 375 (88.7) |
| **Overall: favourable attitude** | **74 (17.5)** |
| **Overall: unfavourable attitude** | **349 (82.5)** |
| **Practices** |  |
| Checked the expiry date before dispensing antibiotics | 307 (72.6) |
| Reported not dispensing antibiotics without a prescription<sup>a</sup> | 15 (3.5) |
| Named at least one antibiotic that they never dispense without a prescription | 187 (44.2) |
| Did not substitute a prescribed antibiotic with another antibiotic<sup>a</sup> | 219 (51.8) |
| Applied different antibiotic dispensing practices for children and adults<sup>a</sup> | 351 (83.0) |
| Returned expired antibiotics to the health zone central office | 45 (10.6) |
| Practised hand hygiene during working hours | 167 (39.5) |
| **Overall: appropriate practice** | **116 (27.4)** |
| **Overall: inappropriate practice** | **307 (72.6)** |

KAP, knowledge, attitudes and practices; AMR, antimicrobial resistance; AWaRe, Access, Watch, Reserve. Values in bold indicate the overall level for each domain, based on the retained items only (knowledge 8, attitudes 9, practices 4 items). “Don’t know” and “prefer not to answer” responses were counted as unfavourable.
<sup>a</sup>Item excluded from the domain score after reliability analysis (Supplementary File S4).

Knowledge of antibiotic use and AMR varied considerably across the items assessed. Most participants correctly recognized that excessive antibiotic use contributes to AMR (82.5%), that self-medication promotes resistance (81.3%), and that inappropriate antibiotic use can worsen disease outcomes (92.0%). However, notable gaps were identified regarding the WHO AWaRe (Access, Watch, Reserve) classification (9.2%), assessment of antimicrobial susceptibility (36.6%), and the pharmacological classification of antibiotics (40.4%). Overall, only 19.4% of participants had good knowledge of antibiotic use and AMR.

Participants’ attitudes also revealed substantial inconsistencies. Although most participants considered regular regulatory inspection of community pharmacies important (85.6%) and agreed that antibiotic treatment should not be discontinued once symptoms improve (88.7%; item excluded from the score), fewer than half expressed attitudes consistent with rational antibiotic dispensing in several common clinical situations. Overall, favourable attitudes toward antibiotic use and AMR were observed in only 74 participants (17.5%). Antibiotic-dispensing practices were similarly poor. Almost all participants (408; 96.5%) reported dispensing antibiotics without a prescription, 232 (56.9%) of them very frequently and 146 (35.8%) usually; only 15 (3.5%) reported not doing so. Fewer than half (187; 44.2%) named at least one antibiotic that they never dispense without a prescription, most often amoxicillin/clavulanate, gentamicin, ceftriaxone or cefixime. Most participants reported checking expiry dates before dispensing (72.6%) and adapting antibiotic dispensing to the patient’s age (83.0%), and 219 (51.8%) reported not substituting a prescribed antibiotic. However, only 45 (10.6%) returned expired antibiotics to the health zone central office; the others reported discarding them with household waste (n = 282), burning them (n = 209) or selling them at a reduced price (n = 9; several answers possible). Only 167 (39.5%) reported practising hand hygiene during working hours. Based on the four retained practice items, only 116 participants (27.4%) were classified as having appropriate antibiotic-dispensing practices.

Taken together, these findings highlight significant gaps across all three KAP domains, with low proportions of participants demonstrating good knowledge (19.4%), favourable attitudes (17.5%), and appropriate practices (27.4%) regarding antibiotic use and AMR.

### Multivariable logistic regression analysis

Table 3 shows the factors independently associated with good knowledge, favourable attitudes and appropriate practices (n = 417). Higher educational attainment was associated with good knowledge (aOR 4.38; 95% CI 1.79–10.74; *P* = 0.001) and with appropriate practices (aOR 5.74; 95% CI 2.55–12.92; *P* < 0.001). Likewise, participants with a pharmaceutical qualification were more likely to have good knowledge (aOR 2.41; 95% CI 1.38–4.20; *P* = 0.002) and appropriate practices (aOR 2.01; 95% CI 1.13–3.58; *P* = 0.018). Sex, professional experience and previous training were not independently associated with good knowledge. Favourable attitudes were independently associated with age (aOR 1.07 per year; 95% CI 1.02–1.11; *P* = 0.002) and with good knowledge (aOR 4.86; 95% CI 2.68–8.82; *P* < 0.001), but not with sex, educational level, professional qualification, experience or previous training. Appropriate practices were also independently associated with age (aOR 1.07 per year; 95% CI 1.03–1.12; *P* < 0.001) and, most strongly, with favourable attitudes (aOR 6.62; 95% CI 3.50–12.53; *P* < 0.001), whereas participants with previous training on antibiotic use and AMR were less likely to report appropriate practices (aOR 0.31; 95% CI 0.18–0.54; *P* < 0.001). Adding health zone to the models did not materially change these estimates (Supplementary File S5). Overall, educational level and pharmaceutical qualification were the main determinants of good knowledge and appropriate practices, good knowledge was associated with favourable attitudes, and favourable attitudes with appropriate dispensing.

**Table 3.** Multivariable logistic regression analysis of factors associated with good knowledge, favourable attitudes and appropriate practices regarding antibiotic use and AMR among community pharmacy workers in Bukavu, Eastern DRC (*n* = 417)

| Variable | Good knowledge |  | Favourable attitudes |  | Appropriate practices |  |
|---|---|---|---|---|---|---|
|  | **aOR (95% CI)** | ***P*** | **aOR (95% CI)** | ***P*** | **aOR (95% CI)** | ***P*** |
| Age (years) | 1.023 (0.983–1.064) | 0.251 | 1.065 (1.024–1.108) | **0.002** | 1.073 (1.032–1.116) | **<0.001** |
| Sex |  |  |  |  |  |  |
| Female | 1.00 (ref.) |  | 1.00 (ref.) |  | 1.00 (ref.) |  |
| Male | 1.532 (0.898–2.615) | 0.117 | 1.353 (0.767–2.388) | 0.297 | 0.659 (0.377–1.152) | 0.144 |
| Highest educational level |  |  |  |  |  |  |
| Primary/secondary | 1.00 (ref.) |  | 1.00 (ref.) |  | 1.00 (ref.) |  |
| University or above | 4.383 (1.790–10.735) | **0.001** | 2.086 (0.911–4.775) | 0.082 | 5.735 (2.546–12.918) | **<0.001** |
| Professional experience (years) | 1.024 (0.966–1.084) | 0.417 | 0.950 (0.888–1.016) | 0.137 | 0.956 (0.893–1.024) | 0.197 |
| Professional qualification |  |  |  |  |  |  |
| Non-pharmaceutical | 1.00 (ref.) |  | 1.00 (ref.) |  | 1.00 (ref.) |  |
| Pharmaceutical | 2.409 (1.380–4.204) | **0.002** | 1.082 (0.572–2.048) | 0.808 | 2.007 (1.125–3.580) | **0.018** |
| Previous training on antibiotic use and AMR |  |  |  |  |  |  |
| No | 1.00 (ref.) |  | 1.00 (ref.) |  | 1.00 (ref.) |  |
| Yes | 1.233 (0.677–2.244) | 0.492 | 0.711 (0.386–1.310) | 0.274 | 0.306 (0.175–0.535) | **<0.001** |
| Knowledge level |  |  |  |  |  |  |
| Poor | – |  | 1.00 (ref.) |  | 1.00 (ref.) |  |
| Good | – |  | 4.859 (2.678–8.817) | **<0.001** | 1.261 (0.660–2.411) | 0.483 |
| Attitude level |  |  |  |  |  |  |
| Unfavourable | – |  | – |  | 1.00 (ref.) |  |
| Favourable | – |  | – |  | 6.618 (3.495–12.532) | **<0.001** |

aOR, adjusted odds ratio; CI, confidence interval; AMR, antimicrobial resistance; ref., reference category. Complete-case analysis (age missing for 6 participants); events: good knowledge 81, favourable attitudes 74, appropriate practices 116. Each model is adjusted for all variables with an estimate in its column; “–” indicates that the variable was not included in that model. Bold values indicate statistical significance (*P* < 0.05).

### Additional findings on “Machange”

A total of 241 (57.0%) of the 423 participants reported being familiar with “Machange”, a locally available mixture of several medicines sold as one preparation. Participants familiar with “Machange” were asked which medicines it usually contains; across all responses, 45 distinct pharmaceutical molecules from several therapeutic classes were reported (Figure 2). Antibiotics were the most frequently reported class (16 molecules; 35.6%), followed by analgesics/non-steroidal anti-inflammatory drugs (NSAIDs; 8; 17.8%), antiparasitic agents (5; 11.1%) and antifungal drugs (4; 8.9%). Of the 236 participants who gave an informative answer on its composition, 235 (99.6%) listed at least one antibiotic. In addition, 158 of the 241 participants familiar with “Machange” (65.6%; 37.4% of all participants) reported selling it. These findings are based on reported formulations and not on laboratory or pharmaceutical verification of the actual composition of “Machange.”

**Figure 2.**
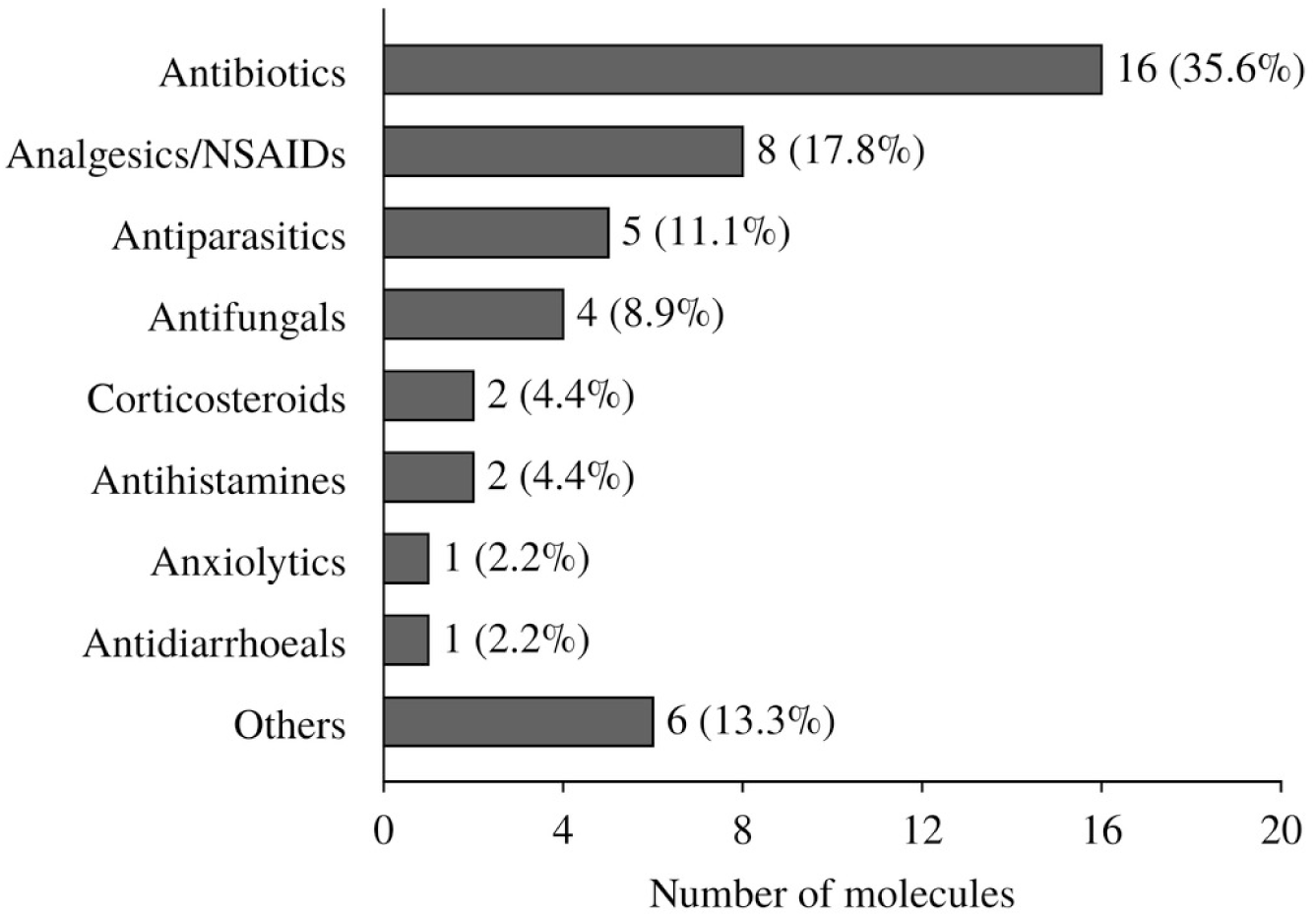
Therapeutic classes of the 45 distinct molecules reported by community pharmacy workers as components of “Machange” preparations. Bars show the number of molecules in each class, with the percentage of the 45 molecules in parentheses. Antiparasitics exclude antimalarials, which are counted under others. Others include vitamins, minerals, antidiabetics, antiemetics, antimalarials and gastrointestinal preparations (one molecule each). NSAIDs, non-steroidal anti-inflammatory drugs.

## Discussion

### Principal findings

Community pharmacy workers play a key role in promoting rational antibiotic use and preventing AMR in LMICs, where pharmacies are often the first point of contact with the health system and are recognized as AMS partners.^2,12,14,29^ As shown in Table 2 and Table 3, community pharmacy workers in Bukavu, Eastern DRC, showed marked deficiencies across all three KAP domains: fewer than one in five had good knowledge (19.4%) or favourable attitudes (17.5%), and only 27.4% reported appropriate dispensing practices, lower than levels reported among qualified pharmacists or in settings with more integrated national AMS programmes.^7,10,13,25,30,31^

Higher education and pharmaceutical qualification were independently associated with better knowledge (four-fold and two-fold increases, respectively), consistent with evidence that academic training predicts AMR-related knowledge,^13,32–34^ while favourable attitudes were the strongest predictor of appropriate practices. This sequential pathway supports the KAP conceptual framework,^13,14^ though educational interventions alone appear insufficient without behavioural-change strategies, regulation, supervision and health-system strengthening.^7,26^

A further contribution was the description of “Machange”, an informal antibiotic-containing drug mixture well known among Bukavu pharmacy workers but not previously documented in the literature. This finding is reinforced by recent microbiological data from Bukavu showing high resistance among clinically important pathogens.^1,3,23,24^ These findings reinforce evidence that education alone cannot resolve stewardship gaps in LMICs; effective stewardship requires clinical training, regulation, supervision, diagnostic capacity and health-system strengthening within a One Health framework, consistent with WHO strategy.^2,6,7,26,35^

### Knowledge regarding antibiotic use and AMR

Overall knowledge of antibiotic use and AMR was low, highlighting gaps among frontline providers and concerns about community-level AMS. Since pharmacies are a principal antibiotic access point in LMICs, this may drive inappropriate dispensing, poor counselling and continued resistance.^2,12^ This is consistent with studies from Jordan,^33^ Palestine,^36^ Ethiopia^17^ and Senegal,^37^ and with a 28-country African survey and a Sri Lankan study reporting widespread deficits despite growing stewardship efforts.^10,13^

The very limited awareness of the WHO AWaRe classification, a central tool for rational antibiotic selection and consumption monitoring,^2,38,39^ echoes reports of limited AWaRe implementation where stewardship and continuing education remain underdeveloped.^7,13,31,40,41^ Limited knowledge of susceptibility testing likely reflects restricted laboratory access and financial barriers common across Africa.^2,3^

These results should be read against a local context of rising AMR amid underdeveloped surveillance and diagnostic capacity.^2,3^ Recent Bukavu microbiological data confirming high resistance among clinically important pathogens show that irrational antibiotic use is a documented, not theoretical, problem.^23,24^ Strengthening pharmacy education is therefore essential, but must be integrated with competency-based training, supervision, stewardship tools and stronger regulation for sustainable improvement.^7,14,26,29^

### Attitudes toward antibiotic use and AMR

Only a small proportion of participants held favourable attitudes towards rational antibiotic use. While many supported routine regulatory inspection and completing antibiotic courses despite symptom improvement, unfavourable attitudes were common regarding non-prescription dispensing and self-limiting conditions. This indicates that knowledge alone does not secure favourable attitudes, which are also shaped by behavioural, regulatory and contextual factors.^26,42,43^

These results echo findings from Ethiopia,^11^ Benin,^44^ Jordan^32^ and Bangladesh,^45^ and align with the 28-country African survey, where higher knowledge did not consistently translate into favourable attitudes or practices.^13^ This suggests the knowledge-attitude-practice gap is a widespread LMIC phenomenon attributable to patient pressure, financial constraints, competition and weak regulatory enforcement.^11,13,44,46^

In multivariable analysis, good knowledge was strongly associated with favourable attitudes (aOR 4.86; 95% CI 2.68–8.82), which were in turn the strongest predictor of good practices (aOR 6.62; 95% CI 3.50–12.53) across all models. Although causality cannot be inferred, this sequential pattern supports the KAP framework and mirrors findings among healthcare professionals elsewhere.^13,14,43,47,48^ Prior antibiotic/AMR training was not independently associated with more favourable attitudes after adjustment, consistent with evidence that education alone rarely produces durable behavioural change without audit, feedback, supervision and multidisciplinary programmes,^7,26^ and with the European Delphi consensus on the need for supportive professional environments and clear guidance.^35^ Building favourable attitudes therefore likely requires strengthening curricula, continuing education, behavioural-change programmes, regulation and integration of pharmacies into national stewardship efforts, rather than knowledge transfer alone.^2,7,12^

### Practices regarding antibiotic use and AMR

Fewer than one-third of participants reported appropriate dispensing practices, with widespread non-prescription dispensing, inappropriate management of expired antibiotics and suboptimal hand hygiene despite reasonable awareness of misuse-related risks, confirming that knowledge does not automatically translate into practice.^2^ Almost all participants dispensed antibiotics without a prescription, and fewer than half restricted even a single antibiotic to prescription-only dispensing. This mirrors evidence that non-prescription dispensing is common in LMIC pharmacies where physician access is limited, consultations are costly and regulation is weak,^25,26^ with comparable findings from Nigeria, South Africa, Sri Lanka, Palestine and Tanzania,^40,49–52^ simulated-patient studies in Jordan,^53^ and the 28-country African survey.^13^

The low rate of appropriate expired-antibiotic disposal, including resale of expired antibiotics by nine participants, is concerning, as improper disposal promotes environmental contamination and selection of resistant organisms, a recognized AMR pathway within the One Health framework.^2,5,6,37,54,55^ Poor hand-hygiene adherence similarly reflects the link between stewardship and infection prevention and control.^2,37^ Unexpectedly, prior training was independently associated with lower odds of appropriate dispensing after adjustment; given the cross-sectional design and lack of data on training content or recency, this may reflect selective training of workers in higher-need settings rather than a causal effect.^7,14,26^ Favourable attitudes remained the strongest independent predictor of good practice (aOR 6.62), mirroring findings among healthcare professionals in Saudi Arabia^48^ and reinforcing that effective stewardship requires continuing education, behavioural change, supervision, regulation, provider-pharmacist collaboration, better diagnostics and stronger health-system governance.^2,7,14,26,35^

### Determinants of KAP

Consistent with the KAP model, higher educational attainment and pharmaceutical qualification were independently associated with better knowledge and practices, mirroring the 28-country African survey^13^ and reflecting greater access to scientific literature and clinical decision-making skills. However, persistent inappropriate practices even among qualified staff indicate that pre-service education alone cannot guarantee rational dispensing, reinforcing the need for continuing education and stewardship-oriented practice environments.^7,14,35^

Age, but not years of experience, was independently associated with attitudes, each additional year raising the odds of favourable attitudes by about 7% (aOR 1.07; 95% CI 1.02–1.11; *P* = 0.002), consistent with KAP studies suggesting clinical maturity, rather than employment duration, shapes stewardship attitudes.^56–58^ The strongest associations overall remained the sequential knowledge-attitude-practice pathway (aOR 4.86 and 6.62, respectively), supporting the KAP model despite the cross-sectional design.^48^ The unexpected inverse association between prior training and good practice likely reflects variation in training quality or timing, residual confounding, or preferential training of higher-risk workers.^7,14,16,26^ These findings support integrated interventions, combining education, continuing development, behavioural change and stronger pharmaceutical governance, within national AMR action plans.^2,7,12,14,35^

### “Machange”: an emerging public health concern

A novel finding was the identification of “Machange”, an informal mixture of pharmaceutical products, predominantly antibiotics alongside analgesics/NSAIDs, antiparasitics and antifungals (Figure 2), known to more than half of participants (57.0%) and sold by about two-thirds of those who knew it. To our knowledge, this is the first report of this practice among community pharmacy workers in the DRC and one of the first in the peer-reviewed literature. Combining multiple antimicrobial and non-antimicrobial products in a single preparation may expose patients to inappropriate antibiotic selection, incorrect dosing, adverse reactions and interactions, and could promote resistance through subtherapeutic exposure and selective pressure, recognized drivers of AMR.^1,2,5^ As this study did not assess its composition, indications or outcomes, no causal relationship with AMR can be established.

This practice likely reflects broader pressures common to resource-limited settings, including poverty, unaffordable healthcare, shortages of prescribers and diagnostics, weak regulation and high out-of-pocket costs, driving informal responses to patients’ immediate needs,^2,7,25^ consistent with adaptive practices documented elsewhere under similar constraints.^14,26^ Given the high proportion of antibiotics reportedly involved, and recent evidence of substantial AMR among clinically important pathogens in Bukavu,^1,2,23,24^ “Machange” warrants urgent attention from researchers, clinicians and public health authorities. Our findings should be regarded as preliminary rather than proof of harm, given the absence of pharmaceutical, microbiological or toxicological data. Future qualitative, mixed-methods and laboratory studies should characterize its composition, motivations for use, economic determinants and contribution to AMR, to inform context-specific stewardship interventions and evidence-based policy in the DRC, and more broadly, informal pharmaceutical practices in resource-limited settings worldwide.

### One Health approach and neglected zoonotic diseases

The One Health approach is particularly relevant to the findings of this study, as AMR transcends human health and involves animal and environmental reservoirs. Neglected zoonotic diseases, such as Q fever, brucellosis, and leptospirosis, are linked to antimicrobial resistance through shared pathways of antibiotic use in human and veterinary medicine, environmental contamination, and inadequate surveillance systems. In resource-limited settings like the DRC, where diagnostic capacity for zoonotic diseases is limited and medical professionals’ knowledge of these conditions is often insufficient,^8^ the One Health approach offers a comprehensive framework for coordinated action. Strategies used in this context include: (i) integrated surveillance systems that monitor AMR across human, animal, and environmental sectors; (ii) responsible antibiotic use in both human and veterinary medicine; (iii) improved infection prevention and control measures, including water, sanitation and hygiene (WASH) interventions; (iv) community education and awareness campaigns targeting both healthcare professionals and the public; and (v) multidisciplinary collaboration between physicians, veterinarians, pharmacists, microbiologists, and public health authorities.^9^ These strategies are essential for addressing the interconnected challenges of AMR and neglected zoonotic diseases, and for preserving the effectiveness of antimicrobial agents for future generations.

### Public health implications

These findings carry important implications for AMS in the DRC and other LMICs. As community pharmacies are among the most accessible entry points into healthcare, suboptimal knowledge, attitudes and dispensing practices may substantially influence antibiotic consumption and the spread of AMR.^2,12,35^ Beyond individual competence, our findings highlight structural barriers, including limited diagnostic and surveillance capacity, workforce shortages, inconsistent stewardship implementation and weak regulation,^1–3^ compounded by high out-of-pocket costs pushing patients toward self-medication through community pharmacies.^20–22,25,59^

Improving stewardship in Bukavu, as elsewhere in sub-Saharan Africa, will require more than educational interventions: stronger regulation, continuing professional development, improved diagnostic and surveillance capacity, and multidisciplinary collaboration between pharmacists, physicians and public health authorities are all needed,^2,7,14,26^ an urgency reinforced by recent evidence of substantial AMR among clinically important pathogens in the DRC.^23,24^ A comprehensive, multisectoral response aligned with the One Health approach, strengthening regulation, education, diagnostic capacity, infection prevention and control, WASH interventions and access to affordable, quality-assured antibiotics, could substantially reduce the future AMR-related mortality burden.^1,2^ Integrating community pharmacies into national AMR action plans, and strengthening the professional, regulatory and health-system environments in which pharmacy workers operate, is therefore a public health priority for the DRC and similar resource-limited settings, in line with the WHO Global Action Plan on AMR.^2,7,12,14,35^

### Strengths and limitations

This study has several strengths. It is among the first to comprehensively assess KAP regarding antibiotic use and AMR among community pharmacy workers in Eastern DRC, addressing an evidence gap in Central Africa, a region underrepresented in the antimicrobial stewardship literature.^3,14^ The sample of 423 workers, recruited across all three health zones of Bukavu using multistage sampling with random selection of pharmacies and workers within each health area, together with a structured questionnaire adapted from previously validated instruments and pre-tested locally, and standardized data collection procedures, supports the reliability of the findings. Multivariable logistic regression, controlling for confounders, further strengthened the internal validity of the identified determinants, moving beyond simple description of KAP levels to inform future interventions and policy. As community pharmacies are increasingly recognized as key AMS partners and are often the first point of access to antibiotics in LMICs,^2,12,29^ these context-specific findings can usefully inform pharmaceutical policy, professional education and stewardship interventions in the DRC. An additional strength was the incidental identification of “Machange” during data collection: although not a pre-specified objective, its repeated mention by participants provided novel evidence of a previously undocumented practice and a starting point for future research from a One Health perspective.

Several limitations should be noted. The cross-sectional design precludes causal inference between the identified determinants and KAP outcomes. Antibiotic dispensing was self-reported and may be affected by recall and social desirability bias, potentially overestimating adherence to appropriate practices. The absence of an up-to-date pharmacy registry in Bukavu prevented a fully probabilistic sampling frame, so residual selection bias cannot be excluded; because one worker was interviewed per pharmacy, workers in larger pharmacies may also be under-represented, and the exclusively urban setting may limit generalizability to rural areas or other DRC provinces. The heterogeneity of the study population, which included both pharmaceutical and non-pharmaceutical personnel, may influence the observed KAP scores and associations; however, this reflects the reality of community pharmacy staffing in the DRC and was accounted for in the analysis through adjustment for professional qualification. Prior antibiotic/AMR training was also self-reported, without verification of its quality, duration or content, which may explain its unexpected association with dispensing practices. The relatively modest Cronbach’s alpha for the practice scale (0.652) after removal of three items should also be acknowledged when interpreting the practice score; because item reduction for the attitude and practice scales was data-driven and the final practice score relies on four items, domain classifications may be sensitive to the choice of items and cut-offs. The cut-off for favourable attitudes (≥60%) was lower than for the other domains because very few participants reached 80%, and the analysis was complete-case (age missing for six participants). Finally, as “Machange” was not a primary study objective, no pharmaceutical, microbiological or toxicological analyses were performed to confirm its composition or effects. Despite these limitations, this study provides an important baseline evidence base on antibiotic-related KAP among community pharmacy workers in Eastern DRC and lays the groundwork for future intervention, operational and One Health research to promote rational antibiotic use and curb AMR in resource-limited settings.

### Recommendations

Findings indicate widespread knowledge gaps (including near-absent awareness of the WHO AWaRe classification), unfavourable attitudes, and inappropriate dispensing (notably non-prescription antibiotic sales, poor management of expired antibiotics, and the emergence of “Machange”), driven largely by weak regulation, an incomplete pharmacy registry, and limited diagnostic capacity. The following recommendations translate these findings into action.

Health authorities should strengthen enforcement of prescription-only dispensing regulations through regular inspections and supportive supervision, establish and maintain an official registry of licensed community pharmacies to close the regulatory gap identified in this study, and formally integrate community pharmacies into the National AMR Action Plan as partners in stewardship, surveillance and public awareness campaigns.^2,12,29^

For stewardship programmes, the low knowledge and AWaRe-awareness observed calls for regular competency-based continuing education on rational antibiotic use, the AWaRe classification, infection prevention and stewardship principles, supported by decision-support tools such as national guidelines and AWaRe-based dispensing algorithms,^2^ and multidisciplinary stewardship networks linking pharmacists, physicians, microbiologists and public health authorities within a One Health approach.^7,14^ At health-system level, addressing the diagnostic and surveillance gaps underlying poor knowledge and inappropriate practice requires investment in microbiological laboratory capacity and antimicrobial susceptibility testing, integration of national AMR surveillance with the WHO Global Antimicrobial Resistance and Use Surveillance System,^2^ reduced financial barriers to formal healthcare to curb self-medication,^25^ and strengthened infection prevention and control and WASH programmes.^1,2^

Professional pharmacy organizations should reinforce mandatory continuing education, certification and competency assessment for dispensing personnel,^12,29^ and strengthen interprofessional collaboration between pharmacists, physicians, nurses and microbiologists.^14,60^ Given the novel and unexplained nature of “Machange”, priority research should characterize its composition, motivations and dispensing patterns through qualitative and mixed-methods studies, and its pharmacological, toxicological and microbiological effects through laboratory-based investigations, alongside implementation research evaluating the cost-effectiveness of community pharmacy stewardship interventions^7,14,26^ and operational and surveillance studies on antibiotic self-medication, financial barriers and AMR trends to inform national policy in the DRC. These research priorities should be implemented within a One Health framework that recognizes the interconnectedness of human, animal and environmental health, and that addresses the structural and feasibility constraints facing resource-limited settings.

## Conclusions

This study provides one of the earliest comprehensive assessments of antibiotic-related KAP among community pharmacy workers in Eastern DRC, revealing substantial deficiencies across all three domains. Better knowledge and dispensing practices were linked to higher education and professional qualification, while favourable attitudes were the strongest predictor of appropriate dispensing, suggesting that effective stewardship may depend on behavioural, regulatory and health-system factors, not knowledge alone. The study also revealed “Machange”, a widely known and widely sold but previously undocumented informal antibiotic mixture requiring urgent further investigation. The near-absent awareness of the WHO AWaRe classification (9.2%) underscores the need for targeted training on this essential stewardship tool. By filling an important evidence gap from a region largely absent from the stewardship literature, these findings can inform pharmaceutical policy, stewardship programmes and future research in the DRC, and reaffirm community pharmacy workers as essential partners in preserving antibiotic effectiveness and combating AMR within a One Health framework.

## Supporting information

Contents Supplementary File S1. STROBE checklist for cross-sectional studies Supplementary File S2. Planned and achieved distribution of surveyed comm

## Declarations

### Ethics statement

The study protocol was reviewed and approved by the Ethics Committee of the Official University of Bukavu (UOB) (approval number UOB/CEM/016/2026). The study was conducted in accordance with the Declaration of Helsinki. Written informed consent was obtained from all participants before each interview, and no personal identifiers were collected. All necessary participant consent has been obtained and the appropriate institutional forms have been archived.

### Competing interests

The authors have declared no competing interest. The authors and their institutions received no payments or services from a third party in the past 36 months that could be perceived to influence, or give the appearance of potentially influencing, the submitted work.

### Funding

This study did not receive any funding. The authors and their institutions received no payment or services from a third party for any aspect of the submitted work.

### Clinical trial registration

Not applicable; this is an observational cross-sectional survey and not a clinical trial.

### Reporting guidelines

This study is reported in accordance with the STROBE guidelines for cross-sectional studies; the completed checklist is provided in Supplementary File S1.

### Author contributions

All authors made a significant contribution to the work reported, whether that is in the conception, study design, execution, acquisition of data, analysis and interpretation, or in all these areas; took part in drafting, revising or critically reviewing the article; gave final approval of this version of the manuscript; and agree to be accountable for all aspects of the work.

Specific contributions: Conceptualization: Willy Mushobekwa Biringanine, David Lupande Mwenebitu. Investigation: Willy Mushobekwa Biringanine, Nancy Rugajo, Justin Ntibonera Aganze, Octave Cito Mudekereza. Data curation: Lameck Zigashane Kawala, Willy Mushobekwa Biringanine. Methodology: Emmanuel Lurhangire Ongezi, Willy Mushobekwa Biringanine, David Lupande Mwenebitu. Formal analysis: Willy Mushobekwa Biringanine. Writing – original draft: Willy Mushobekwa Biringanine. Writing – review and editing: David Lupande Mwenebitu, Willy Mushobekwa Biringanine, Corneille Kabagale Ansima. Supervision: David Lupande Mwenebitu.

### Data availability

The datasets generated and/or analysed during the current study are available from the corresponding author upon reasonable request. The questionnaire and scoring key, the planned and achieved sample distribution by health area, the item-level reliability statistics, the bivariate analyses and the STROBE checklist are provided as Supplementary Files S1–S5.

### Supplementary material

Supplementary Files S1–S5 are provided as a separate supplementary file accompanying this preprint.

## Acknowledgements

The authors used ChatGPT (GPT-5, OpenAI, San Francisco, USA) and Claude (Claude Sonnet 5, Anthropic, San Francisco, USA) to improve the English language, coherence and formatting of this manuscript. All scientific content, interpretations and conclusions remain the sole responsibility of the authors.

## Notes

### Author Declarations

Ethics committee/IRB of Universite officielle de Bukavu gave ethical approval for this work

## References

1. Naghavi M, Vollset SE, Ikuta KS et al. Global burden of bacterial antimicrobial resistance 1990–2021: a systematic analysis with forecasts to 2050. Lancet 2024; 404: 1199–226.

2. WHO. Global Antibiotic Resistance Surveillance Report 2025. 2025. https://www.who.int/publications/i/item/9789240116337

3. Tadesse BT, Ashley EA, Ongarello S et al. Antimicrobial resistance in Africa: a systematic review. BMC Infect Dis 2017; 17: 616.

4. O’Neill J. Tackling Drug-Resistant Infections Globally: Final Report and Recommendations. Review on Antimicrobial Resistance, 2016.

5. Yao J, Zou P, Cui Y et al. Recent advances in strategies to combat bacterial drug resistance: antimicrobial materials and drug delivery systems. Pharmaceutics 2023; 15: 1188.

6. Holmes AH, Moore LSP, Sundsfjord A et al. Understanding the mechanisms and drivers of antimicrobial resistance. Lancet 2016; 387: 176–87.

7. Kakkar P. Community antimicrobial stewardship: what actually works? (Education, audit-feedback, dispensing interventions) — a systematic review focused on UK and low- and middle-income country (LMIC) settings. Int J Innov Sci Res Technol 2025; 10: 662–79.

8. Meurer IR, Silva MR, Roland RK et al. Evaluation of medical professionals’ knowledge about Q fever. Sci Med 2024; 34: e45474.

9. Meurer IR. The importance of medical knowledge about Q fever in the context of timely diagnosis and treatment and the use of the One Health approach in combating this and other neglected zoonotic diseases. Infect Drug Resist 2025; 18: 5007–8.

10. Sakeena MHF, Bennett AA, McLachlan AJ. Non-prescription sales of antimicrobial agents at community pharmacies in developing countries: a systematic review. Int J Antimicrob Agents 2018; 52: 771–82.

11. Belachew SA, Hall L, Selvey LA. Community drug retail outlet staff’s knowledge, attitudes and practices towards non-prescription antibiotics use and antibiotic resistance in the Amhara region, Ethiopia with a focus on non-urban towns. Antimicrob Resist Infect Control 2022; 11: 64.

12. International Pharmaceutical Federation. FIP Statement of Policy: Mitigating Antimicrobial Resistance (AMR) Through Antimicrobial Stewardship (AMS). FIP, 2023.

13. Agiri O, Osena G, Bahati F et al. Knowledge, attitudes and practices survey on antimicrobial resistance and stewardship among pharmacy healthcare workers in 28 African countries. BMJ Glob Health 2025; 10: e019151.

14. Saha SK, Barton C, Promite S et al. Knowledge, perceptions and practices of community pharmacists towards antimicrobial stewardship: a systematic scoping review. Antibiotics (Basel) 2019; 8: 263.

15. Kariuki S. Global burden of antimicrobial resistance and forecasts to 2050. Lancet 2024; 404: 1172–3.

16. Raju R, Srinivas SC, Siddalingegowda SM et al. Community pharmacists as antimicrobial resistance stewards: a narrative review on their contributions and challenges in low- and middle-income countries. J Pharm Pharm Sci 2024; 27: 12721.

17. Erku DA, Mersha AG. Involvement of community pharmacists in public health priorities: a multi-center descriptive survey in Ethiopia. PLoS One 2017; 12: e0180943.

18. Musoke D, Lubega GB, Gbadesire MS et al. Antimicrobial stewardship in private pharmacies in Wakiso district, Uganda: a qualitative study. J Pharm Policy Pract 2023; 16: 147.

19. Lupande-Mwenebitu D, Baron SA, Nabti LZ et al. Current status of resistance to antibiotics in the Democratic Republic of the Congo: a review. J Glob Antimicrob Resist 2020; 22: 818–25.

20. Maini R, Van den Bergh R, van Griensven J, et al. Picking up the bill—improving health-care utilisation in the Democratic Republic of Congo through user fee subsidisation: a before and after study. BMC Health Serv Res 2014; 14: 504.

21. Bapolisi WA, Karemere H, Ndogozi F et al. First recourse for care-seeking and associated factors among rural populations in the eastern Democratic Republic of the Congo. BMC Public Health 2021; 21: 1367.

22. Hermes Karemere., et al. “Analysis of the Factors Limiting Access to Health Services in the Kalehe Health Zone in the Democratic Republic of Congo". Acta Scientific Medical Sciences 9.6 (2025): 137–145.

23. Cito B, Corneille K, Gates I et al. Bacteriological profile and antibiotic resistance patterns of uropathogenic bacteria at HPGRB. Infect Drug Resist 2026; 19: 1–14.

24. Ansima CK, Mulume GI, Maroyi BJ et al. High Burden of Multidrug-Resistant Bacteria in Eastern Democratic Republic of the Congo: A Retrospective Hospital-Based Study. Infect Drug Resist. 2026;19:626233

25. Auta A, Hadi MA, Oga E et al. Global access to antibiotics without prescription in community pharmacies: a systematic review and meta-analysis. J Infect 2019; 78: 8–18.

26. Thandar MM, Baba T, Matsuoka S et al. Interventions to reduce non-prescription antimicrobial sales in community pharmacies. Cochrane Database Syst Rev 2025; issue 1: CD013722.

27. Bahinzi LZM, Bepouka B, Mudogo CN et al. Knowledge, attitudes and practices of healthcare providers on antibiotics use and resistance in hospital settings in Kinshasa, Democratic Republic of Congo. Infect Drug Resist 2025; 18: 4023–37.

28. Shrestha B, Dunn L. The Declaration of Helsinki on medical research involving human subjects: a review of seventh revision. J Nepal Health Res Counc 2019; 17: 548–52.

29. Lambert M, van Dijk L, Benko R et al. The role of the community pharmacist in antibiotic use— a commentary on current status and future perspectives. Explor Res Clin Soc Pharm 2025; 20: 100652.

30. Fenech B, Gaffiero D. Barriers and facilitators to antimicrobial stewardship in antibiotic prescribing and dispensing by general practitioners and pharmacists in Malta: a systematic review. Antibiotics (Basel) 2025; 14: 1181.

31. Plaza Zamora FJ, Mendoza Barbero A, Molinero A, et al. Second audit of the Happy Patient project: impact of training on the dispensing of antibiotics in community pharmacies in Spain. Farm Comunitarios 2026; 18: e249.

32. Darwish RM, Baqain GN, Aladwan H et al. Knowledge, attitudes, and practices regarding antibiotic use and resistance among community pharmacists: a cross sectional study in Jordan. Int J Clin Pharm 2021; 43: 1198–207.

33. Abdel-Qader DH, Albassam A, Ismael NS et al. Community pharmacists’ knowledge of and attitudes toward antibiotic use, resistance, and self-medication in Jordan. Drugs Ther Perspect 2021; 37: 44–53.

34. Salah AN, Aboelyazed AM, Waseem AM et al. The practice and knowledge among community pharmacists about dispensing antibiotics in Egypt; a cross-sectional study. Microbes Infect Dis 2025; 6(2): 575–587

35. Lambert M, Wonink A, Benko R et al. Pharmacists combating antimicrobial resistance: a Delphi study on antibiotic dispensing. Res Social Adm Pharm 2024; 20: 723–32.

36. Aiesh BM, Saleh S, Matouk N et al. Assessing pediatric antibiotic knowledge and practices among community pharmacists in Palestine: implications for antibiotic use and resistance. BMC Pediatr 2024; 24: 582.

37. Oumar O, Awa BD. Community pharmacists’ knowledge, practices and perceptions on antibiotic use and resistance: a cross-sectional, self-administered questionnaire survey, in Guediawaye and Pikine, Senegal. Open J Epidemiol 2019; 9: 289–308.

38. Hsia Y, Lee BR, Versporten A et al. Use of the WHO Access, Watch, and Reserve classification to define patterns of hospital antibiotic use (AWaRe): an analysis of paediatric survey data from 56 countries. Lancet Glob Health 2019; 7: e861–71.

39. Pauwels I, Versporten A, Drapier N et al. Hospital antibiotic prescribing patterns in adult patients according to the WHO Access, Watch and Reserve classification (AWaRe): results from a worldwide point prevalence survey in 69 countries. J Antimicrob Chemother 2021; 76: 1614–24.

40. Maluleke TM, Maluleke MT, Jelic AG et al. Estimated extent of purchasing of antibiotics without a prescription from community pharmacies in a rural province in South Africa and the implications. Front Trop. Dis. 6:1637362.

41. Sultana Samapti MM, Hossain KM, Riha FA et al. Understanding the role of knowledge and attitudes of community pharmacy staff in non-prescribed antibiotic dispensing practices in Bangladesh: a cross-sectional study. BMJ Open 2026; 16: e107698.

42. Bishop C, Yacob Z, Knobloch MJ et al. Community pharmacy interventions to improve antibiotic stewardship and implications for pharmacy education: a narrative overview. Res Social Adm Pharm 2019; 15: 627–31.

43. Zerbinato F, Cunningham S, Tonna AP. Antimicrobial stewardship interventions involving community pharmacy teams: a scoping review. JAC Antimicrob Resist 2025; 7: dlaf156.

44. Allabi AC, Agbo AG, Boya B et al. Antimicrobial stewardship: knowledge and attitudes of pharmacy staff on antibiotic dispensing patterns, use and resistance in Benin. Pharmacol Pharm 2023; 14: 189–214.

45. Al Masud A, Walpola RL, Sarker M et al. Assessing community antibiotic usage and adherence as per standard treatment guidelines: a potential area to enhance awareness at community pharmacy settings. Explor Res Clin Soc Pharm 2025; 17: 100552.

46. Miner TW, Callaway Kim K, Rothenberger S et al. Impact of implementation of World Health Organization National Action Plans on antibiotic rates: a time series analysis of 37 countries. Infect Control Hosp Epidemiol 2025; 46: 1171–4.

47. Jamshed S, Padzil F, Shamsudin SH et al. Antibiotic stewardship in community pharmacies: a scoping review. Pharmacy (Basel) 2018; 6: 92.

48. Alajmi AM, Alamoudi AA, Halwani AA et al. Antimicrobial resistance awareness, antibiotics prescription errors and dispensing patterns by community pharmacists in Saudi Arabia. J Infect Public Health 2023; 16: 34–41.

49. Akande-Sholabi W, Oyesiji E, Adebisi YA. Antimicrobial stewardship: community pharmacists’ antibiotic dispensing practices, knowledge, and perception regarding antibiotics and antibiotic resistance. J Pharm Health Serv Res 2023; 14: 383–91.

50. Zawahir S, Lekamwasam S, Aslani P. A cross-sectional national survey of community pharmacy staff: knowledge and antibiotic provision. PLoS One 2019; 14: e0215484.

51. Al-Halawa DA, Seir RA, Qasrawi R. Antibiotic resistance knowledge, attitudes, and practices among pharmacists: a cross-sectional study in West Bank, Palestine. J Environ Public Health 2023; 2023: 2294048.

52. Horumpende PG, Sonda TB, van Zwetselaar M et al. Prescription and non-prescription antibiotic dispensing practices in part I and part II pharmacies in Moshi Municipality, Kilimanjaro Region in Tanzania: a simulated clients approach. PLoS One 2018; 13: e0207465.

53. Jarab AS, Al-Qerem W, Alzoubi KH et al. Pharmacists’ knowledge, attitudes, and practices of dispensing antibiotics without a prescription. Pharm Pract (Granada) 2025; 23: 1–8.

54. Mshana SE, Matee M, Rweyemamu M. Antimicrobial resistance in human and animal pathogens in Zambia, Democratic Republic of Congo, Mozambique and Tanzania: an urgent need of a sustainable surveillance system. Ann Clin Microbiol Antimicrob 2013; 12: 28.

55. Collineau L, Colomb-Cotinat M. L’approche une seule santé pour une action coordonnée contre l’antibiorésistance—enjeux et défis. Med Sci (Paris) 2026; 42: 175–83.

56. Abubakar U, Tangiisuran B. Knowledge and practices of community pharmacists towards non-prescription dispensing of antibiotics in Northern Nigeria. Int J Clin Pharm 2020; 42: 756–64.

57. Khan MU, Hassali MAA, Ahmad A et al. Perceptions and practices of community pharmacists towards antimicrobial stewardship in the State of Selangor, Malaysia. PLoS One 2016; 11: e0149623.

58. Nabeel M, Ali K, Sarwar MR et al. Assessment of knowledge, attitudes, and practices among community pharmacists in Lahore regarding antibiotic dispensing without prescription: a cross-sectional study. PLoS One 2024; 19: e0304361.

59. Shrestha N, Manandhar S, Maharjan N et al. Perspectives of pharmacy employees on an inappropriate use of antimicrobials in Kathmandu, Nepal. PLoS One 2023; 18: e0285287.

60. Kakati R, Nakad Borrego S, Zareef R et al. Dispensing and purchasing antibiotics without prescription: a cross-sectional study among pharmacists and patients in Beirut, Lebanon. Inquiry 2023; 60: 469580231167712.

