## Supplementary material for "Knowledge, attitudes and practices regarding antibiotic use and antimicrobial resistance among community pharmacy workers in Bukavu, Eastern Democratic Republic of Congo: a cross-sectional study": Contents Supplementary File S1. STROBE checklist for cross-sectional studies Supplementary File S2. Planned and achieved distribution of surveyed comm

Supplementary File S1. STROBE checklist for cross-sectional studies

Supplementary File S2. Planned and achieved distribution of surveyed community pharmacies by health area

Supplementary File S3. Study questionnaire and scoring key

Supplementary File S4. Item-level reliability analysis of the knowledge, attitude and practice scales

Supplementary File S5. Bivariate associations and sensitivity analysis

### Supplementary File S1. STROBE checklist for cross-sectional studies

Knowledge, attitudes and practices regarding antibiotic use and antimicrobial resistance among community pharmacy workers in Bukavu, Eastern Democratic Republic of Congo: a cross-sectional study

Locations refer to the section headings of the main manuscript.

| Item | No. | Recommendation | Where reported |
|---|---|---|---|
| Title and abstract | 1 | (a) Design in title/abstract; (b) informative, balanced summary | Title ("cross-sectional study"); Abstract |
| Background/rationale | 2 | Scientific background and rationale | Introduction, paragraphs 1–5 |
| Objectives | 3 | Specific objectives | Introduction, final paragraph |
| Study design | 4 | Key elements of design early in the paper | Methods – Study design |
| Setting | 5 | Setting, locations, dates of recruitment and data collection | Methods – Study setting; Data collection procedures (April–June 2026); Figure 1 |
| Participants | 6 | Eligibility criteria, sources and methods of selection | Methods – Study population; Eligibility criteria; Sampling procedure; Supplementary File S2 |
| Variables | 7 | Outcomes, exposures, potential confounders | Methods – Study variables; Assessment of KAP; Statistical analysis |
| Data sources/measurement | 8 | Sources and methods of assessment | Methods – Data collection instrument; Assessment of KAP; Assessment of internal consistency; Supplementary Files S3, S4 |
| Bias | 9 | Efforts to address potential sources of bias | Methods – Sampling procedure (random selection of pharmacies and workers); Data collection procedures (trained interviewers, private setting); Discussion – Strengths and limitations |
| Study size | 10 | How study size was arrived at | Methods – Sample size determination |
| Quantitative variables | 11 | Handling of quantitative variables, groupings | Methods – Study variables (dichotomization); Statistical analysis (age and experience kept continuous) |
| Statistical methods | 12 | (a) Methods incl. confounding control; (b) subgroups/interactions; (c) missing data; (d) sampling strategy; (e) sensitivity analyses | (a) Statistical analysis; (b) not performed; (c) Statistical analysis (complete-case; age missing for 6 participants); (d) analyses not weighted for the multistage design; (e) sensitivity analysis adding health zone (Supplementary File S5) |
| Participants | 13 | Numbers at each stage; reasons for non-participation; flow diagram | Methods – Sample size determination (100% response) and Sampling procedure; Supplementary File S2; flow diagram not provided |
| Descriptive data | 14 | Characteristics of participants; missing data | Results; Table 1 (footnote: age missing for 6 participants) |
| Outcome data | 15 | Numbers of outcome events | Results; Table 2 |
| Main results | 16 | Unadjusted and adjusted estimates with precision; category boundaries | Results – Multivariable logistic regression; Table 3; Supplementary File S5 (crude estimates); Methods – Assessment of KAP (cut-offs) |
| Other analyses | 17 | Other analyses | Results – Additional findings on "Machange"; Figure 2 |

| Item | No. | Recommendation | Where reported |
|---|---|---|---|
| Key results | 18 | Summary of key results | Discussion – Principal findings |
| Limitations | 19 | Limitations, sources of bias or imprecision | Discussion – Strengths and limitations |
| Interpretation | 20 | Cautious overall interpretation | Discussion, including Conclusions |
| Generalizability | 21 | External validity | Discussion – Strengths and limitations |
| Funding | 22 | Source of funding and role of funders | Declarations – Funding |

### Supplementary File S2. Planned and achieved distribution of surveyed community pharmacies by health area

Knowledge, attitudes and practices regarding antibiotic use and antimicrobial resistance among community pharmacy workers in Bukavu, Eastern Democratic Republic of Congo: a cross-sectional study

The target of 423 pharmacies was allocated to health areas in proportion to their population (2025 South Kivu Provincial Health Report). In each health area, pharmacies were selected at random from an on-site list of operating pharmacies, and one worker was interviewed per pharmacy, so achieved numbers of pharmacies equal numbers of participants. Health-area names are those recorded in the study database; some differ from the labels on the map in Figure 1.

| Health zone | Health area | Population (2025) | Planned (n) | Achieved (n) |
|---|---|---|---|---|
| **Ibanda health zone (17 health areas with pharmacies)** | | | | |
| Ibanda | Ceca40 | 31697 | 8 | 10 |
| Ibanda | Chahi | 38907 | 10 | 7 |
| Ibanda | Cidasa | 62779 | 16 | 12 |
| Ibanda | Cr Nguba | 33815 | 9 | 10 |
| Ibanda | Cr Saio | 24064 | 6 | 4 |
| Ibanda | Gihamba | 29241 | 8 | 7 |
| Ibanda | Ibanda | 16028 | 4 | 10 |
| Ibanda | Irambo | 18163 | 5 | 5 |
| Ibanda | Kabuye | 23243 | 6 | 0 |
| Ibanda | Malkia | 26628 | 7 | 4 |
| Ibanda | Maman Mwilu | 62741 | 16 | 11 |
| Ibanda | Muhungu Dioc | 23847 | 6 | 8 |
| Ibanda | Muhungu Etat | 46893 | 12 | 14 |
| Ibanda | Mulungulungu | 16772 | 4 | 14 |
| Ibanda | Nyawera | 22869 | 6 | 8 |
| Ibanda | Panzi | 41546 | 11 | 7 |
| Ibanda | Vavassori | 28017 | 7 | 15 |
| Ibanda | Subtotal | 547248 | 141 | 146 |
| **Kadutu health zone (13 health areas with pharmacies)** | | | | |
| Kadutu | Biname | 37664 | 15 | 8 |
| Kadutu | Buholo 2 | 32801 | 13 | 20 |
| Kadutu | Ceca Mweze | 33567 | 14 | 16 |
| Kadutu | Cimpunda | 35069 | 14 | 14 |
| Kadutu | Ciriri 1 | 62466 | 26 | 21 |
| Kadutu | Ciriri 2 | 23609 | 10 | 14 |
| Kadutu | Funu | 27181 | 11 | 9 |
| Kadutu | Lurhuma | 22171 | 9 | 6 |
| Kadutu | Mariia/Karhale | 81635 | 34 | 29 |
| Kadutu | Neema | 25135 | 10 | 14 |
| Kadutu | Nyamugo | 30737 | 13 | 9 |
| Kadutu | Nyamulagira | 26303 | 11 | 3 |
| Kadutu | Uzima | 17323 | 7 | 19 |
| Kadutu | Subtotal | 455662 | 187 | 182 |

| Health zone | Health area | Population (2025) | Planned (n) | Achieved (n) |
|---|---|---|---|---|
| **Bagira health zone (6 health areas with pharmacies)** | | | | |
| Bagira | Bagira | 18259 | 10 | 17 |
| Bagira | Burhiba | 34545 | 19 | 40 |
| Bagira | Kaheru | 17687 | 11 | 12 |
| Bagira | Lumu | 22017 | 12 | 7 |
| Bagira | Makoma | 14915 | 8 | 7 |
| Bagira | Nyamuhinga | 23826 | 13 | 12 |
| **Health areas without an operating community pharmacy** | | | | |
| Bagira | Ciguri | 11374 | 6 | 0 |
| Bagira | Mushekere | 29440 | 16 | 0 |
| Bagira | Subtotal | 172063 | 95 | 95 |
| Bukavu | Total (38 health areas) | **1174973** | **423** | **423** |

Two health areas (Ciguri and Mushekere) had no operating community pharmacy at the time of the survey; their planned allocation was reallocated as described in the Methods.

### Supplementary File S3. Study questionnaire and scoring key

Knowledge, attitudes and practices regarding antibiotic use and antimicrobial resistance among community pharmacy workers in Bukavu, Eastern Democratic Republic of Congo: a cross-sectional study

Interviewer-administered in French with KoboCollect, April–June 2026; one worker per selected community pharmacy; written informed consent obtained before the interview. The dataset variable corresponding to each item is given in the last column of the scoring tables.

#### Section A. Sociodemographic and professional characteristics

| No. | Question | Response options |
|---|---|---|
| Q1 | Age (completed years) | Number |
| Q2 | Sex | Female / Male |
| Q3 | Highest level of education completed | Primary / Secondary / University / Postgraduate (specialization) |
| Q4 | Profession / professional qualification | Pharmacist / Pharmacy technician (G3) / Pharmacy assistant (A2) / Physician / Nurse / Laboratory technician / Midwife / Other (specify) |
| Q5 | Years of work experience in the pharmacy | Number |
| Q6 | Health zone of the pharmacy | Ibanda / Kadutu / Bagira |
| Q7 | Health area of the pharmacy | List of health areas |
| Q8 | Have you received training on antibiotic use and antimicrobial resistance? | Yes / No |
| Q9 | What are your sources of information on antibiotics? (several answers possible) | Pharmacology course / Package inserts / Internet / Pharmaceutical sales representative / Antibiotic guidelines and leaflets / Other |

#### Section B. Knowledge (8 items; score 1 = correct answer, 0 otherwise)

| No. | Question | Response options | Scored 1 | Variable |
|---|---|---|---|---|
| K1 | Have you heard of the WHO AWaRe (Access, Watch, Reserve) classification of antibiotics? | Yes / No | Yes | knc_aware |
| K2 | Can the (excessive) use of antibiotics lead to antimicrobial resistance? | Yes / No / Don't know | Yes | usag_atb_ram |
| K3 | Does antimicrobial resistance make infections more difficult to treat? | Yes / No / Don't know | Yes | ram_t3_dificil |
| K4 | Do you know what an antibiogram (antimicrobial susceptibility test) is and what it is used for? | Yes / No | Yes | knc_atbgm |
| K5 | Does self-medication with antibiotics contribute to antimicrobial resistance in the DRC? | Yes / No / Don't know | Yes | automed_ram_rdc |
| K6 | Is hand hygiene an effective measure to prevent infections? | Yes / No / Don't know | Yes | hygiene_main_infectio |
| K7 | Can taking antibiotics inappropriately worsen a disease? | Yes / No / Don't know | Yes | mauv_prise_atb_maladi |
| K8 | Do you know the pharmacological classes of antibiotics? | Yes / No | Yes | knc_class_atb |

#### Section C. Attitudes (10 items; score 1 = favourable answer, 0 otherwise)

| No. | Question | Response options | Scored 1 | Variable |
|---|---|---|---|---|
| A1 | In the past 3 months, have you proposed antibiotics to clients? | Yes / No / Don't know | No | prop_atb_3m |
| A2 | Should antibiotics be dispensed only on presentation of a prescription? | Yes / No / Don't know | Yes | ordonance_atb |
| A3 | Is personal experience sufficient to dispense an antibiotic? | Yes / No / Don't know | No | atb_experience |
| A4 | Would you dispense an antibiotic for fever, cold or cough lasting less than 2 days? | Yes / No / Don't know | No | atb_f_r_t |
| A5 | Would you dispense an antibiotic for acute diarrhoea lasting less than 1 day? | Yes / No / Don't know | No | atb_diarrhee |
| A6 | Would you dispense an antibiotic for a simple cough? | Yes / No / Don't know | No | atb_toux |
| A7 | Which antibiotic would you dispense for influenza (flu)? | Free text (name of antibiotic, or "no indication") | No antibiotic indicated | atb_grippe |
| A8 | Do you advise clients to consult a doctor before taking antibiotics? | Yes / No / Prefer not to answer | Yes | conseil_consul_med |
| A9 | Do you think that regular inspection of community pharmacies by the authorities is important? | Yes / No / Don't know | Yes | importance_controle |
| A10 | Should a patient continue the antibiotic even after feeling better? | Yes / No / Don't know | Yes | atb_sentir_bien |

#### Section D. Antibiotic-dispensing practices (7 items, self-reported; score 1 = appropriate answer, 0 otherwise)

| No. | Question | Response options | Scored 1 | Variable |
|---|---|---|---|---|
| P1 | Do you check the expiry date before dispensing an antibiotic? | Yes / No | Yes | date_per_verification |
| P2 | Do you dispense antibiotics without a prescription? If yes, how often? | Yes (very frequently / usually / less frequently) / No | No | atb_sans_ord |
| P3 | Are there antibiotics that you never dispense without a prescription? If yes, which? | Yes (specify) / No | Yes | atb_jamais_s_ord |
| P4 | Do you propose an antibiotic other than the one prescribed? If yes, why? | Yes (client request / based on training / availability in the pharmacy) / No / Prefer not to answer | No | proposition_atb_ord |
| P5 | Do you dispense antibiotics in the same way to children and adults? If not, on what basis? | Yes / No (age, weight, dose) | No | atb_eft_ad |
| P6 | What do you do with expired antibiotics? (several answers possible) | Throw in the waste bin / Burn / Return to the health zone central office (BCZS) / Sell at a low price to clients in difficulty | Return to BCZS only | atb_perimes |
| P7 | Do you wash your hands during working hours? | Yes / Not very often / No | Yes | hygiene_main_h_servic |

### Section E. Other items collected (not part of the KAP scores)

| No. | Question | Response options |
|---|---|---|
| E1 | Do you perform a physical examination before dispensing an antibiotic? If yes, which? | Yes (inspection / palpation) / No / Prefer not to answer |
| E2 | Have you ever encountered falsified expiry dates on medicines? | Yes / No |
| E3 | Has your pharmacy been inspected by the Provincial Health Inspectorate? If yes, how many times? | Yes (number) / No / Don't know |
| E4 | What problems, if any, have you encountered during inspections? | Free text |

### Section F. “Machange”

| No. | Question | Response options |
|---|---|---|
| M1 | Do you know the preparation called "Machange"? | Yes / No |
| M2 | If yes: what does "Machange" usually contain? (list the medicines) | Free text, coded afterwards into molecules and therapeutic classes |
| M3 | If yes: do you sell "Machange"? | Yes / No |

### Scoring key

| Domain | Items in final score | Cut-off | Classification |
|---|---|---|---|
| Knowledge | K1–K8 (8 items) | ≥80% of maximum (≥7/8) | Good vs poor knowledge |
| Attitudes | A1–A9 (9 items; A10 excluded) | ≥60% of maximum (≥6/9) | Favourable vs unfavourable attitude |
| Practices | P1, P3, P6, P7 (4 items; P2, P4, P5 excluded) | ≥75% of maximum (≥3/4) | Appropriate vs inappropriate practice |

"Don't know" and "prefer not to answer" were scored 0. Excluded items (Supplementary File S4) are reported descriptively in Table 2 of the main text.

### Supplementary File S4. Item-level reliability analysis of the knowledge, attitude and practice scales

Knowledge, attitudes and practices regarding antibiotic use and antimicrobial resistance among community pharmacy workers in Bukavu, Eastern Democratic Republic of Congo: a cross-sectional study

Cronbach's alpha was calculated for each initial scale. Items with a corrected item–total (item–rest) correlation below 0.10 were excluded from the final domain score. n (%) is the number of participants giving the correct, favourable or appropriate response (N = 423). Statistics refer to the initial scales.

| Item | Description | n (%) | Corrected item–total correlation | Alpha if item deleted | Retained in score |
|---|---|---|---|---|---|
| **Knowledge — initial alpha 0.707 (8 items); final alpha 0.707 (8 items)** | | | | | |
| K1 | Awareness of the WHO AWaRe classification | 39 (9.2) | 0.249 | 0.705 | Yes |
| K2 | Recognized excessive antibiotic use as a driver of AMR | 349 (82.5) | 0.557 | 0.644 | Yes |
| K3 | Recognized that AMR can make infections difficult to treat | 338 (79.9) | 0.520 | 0.651 | Yes |
| K4 | Knowledge of antimicrobial susceptibility testing | 155 (36.6) | 0.439 | 0.670 | Yes |
| K5 | Recognized self-medication as a driver of AMR | 344 (81.3) | 0.533 | 0.649 | Yes |
| K6 | Recognized hand hygiene as effective | 315 (74.5) | 0.215 | 0.720 | Yes |
| K7 | Recognized inappropriate use may worsen disease outcomes | 389 (92.0) | 0.374 | 0.687 | Yes |
| K8 | Knowledge of the pharmacological classification of antibiotics | 171 (40.4) | 0.363 | 0.690 | Yes |
| **Attitudes — initial alpha 0.692 (10 items); final alpha 0.710 (9 items)** | | | | | |
| A1 | Did not routinely recommend antibiotics to clients | 7 (1.7) | 0.139 | 0.696 | Yes |
| A2 | Agreed antibiotics should only be dispensed with a prescription | 171 (40.4) | 0.422 | 0.654 | Yes |
| A3 | Disagreed that personal experience alone is sufficient | 91 (21.5) | 0.186 | 0.697 | Yes |
| A4 | Did not support antibiotics for fever, cold or cough <2 days | 164 (38.8) | 0.413 | 0.656 | Yes |
| A5 | Did not support antibiotics for acute diarrhoea <1 day | 121 (28.6) | 0.531 | 0.632 | Yes |
| A6 | Did not support antibiotics for cough alone | 94 (22.2) | 0.512 | 0.639 | Yes |
| A7 | Recognized influenza does not routinely require antibiotics | 132 (31.2) | 0.498 | 0.638 | Yes |
| A8 | Recommended medical consultation before antibiotic use | 190 (44.9) | 0.481 | 0.641 | Yes |
| A9 | Considered regular regulatory inspections important | 362 (85.6) | 0.174 | 0.696 | Yes |
| A10 | Agreed treatment should not be stopped once symptoms improve | 375 (88.7) | 0.050 | 0.710 | No |
| **Practices — initial alpha 0.521 (7 items); final alpha 0.652 (4 items)** | | | | | |
| P1 | Checked the expiry date before dispensing | 307 (72.6) | 0.446 | 0.391 | Yes |
| P2 | Reported not dispensing antibiotics without a prescription | 15 (3.5) | 0.059 | 0.535 | No |

| Item | Description | n (%) | Corrected item–total correlation | Alpha if item deleted | Retained in score |
|---|---|---|---|---|---|
| P3 | Named at least one antibiotic never dispensed without a prescription | 187 (44.2) | 0.422 | 0.396 | Yes |
| P4 | Did not substitute a prescribed antibiotic | 219 (51.8) | 0.088 | 0.566 | No |
| P5 | Applied different dispensing for children and adults | 351 (83.0) | 0.090 | 0.543 | No |
| P6 | Returned expired antibiotics to the health zone central office | 45 (10.6) | 0.255 | 0.487 | Yes |
| P7 | Practised hand hygiene during working hours | 167 (39.5) | 0.424 | 0.395 | Yes |

### Supplementary File S5. Bivariate associations and sensitivity analysis

Knowledge, attitudes and practices regarding antibiotic use and antimicrobial resistance among community pharmacy workers in Bukavu, Eastern Democratic Republic of Congo: a cross-sectional study

**Table A.** Crude associations between candidate explanatory variables and good knowledge, favourable attitudes and appropriate practices (N = 423; n = 417 for age). Crude odds ratios from univariable logistic regression. P values: Pearson chi-square (Fisher exact test when an expected count was <5) for categorical variables, with one global test for health zone; Wilcoxon rank-sum test for age and experience.

| Variable | Good knowledge n/N (%) | Crude OR (95% CI) | *P* | Favourable attitude n/N (%) | Crude OR (95% CI) | *P* | Appropriate practice n/N (%) | Crude OR (95% CI) | *P* |
|---|---|---|---|---|---|---|---|---|---|
| Age (per year) | | 1.04 (1.01–1.06) | 0.015 | | 1.05 (1.03–1.08) | <0.001 | | 1.05 (1.03–1.08) | <0.001 |
| **Sex** | | | | | | | | | |
| Female (ref.) | 43/275 (15.6) | 1.00 | | 40/275 (14.5) | 1.00 | | 77/275 (28.0) | 1.00 | |
| Male | 39/148 (26.4) | 1.93 (1.18–3.15) | 0.008 | 34/148 (23.0) | 1.75 (1.05–2.92) | 0.030 | 39/148 (26.4) | 0.92 (0.59–1.44) | 0.717 |
| **Highest educational level** | | | | | | | | | |
| Primary/secondary (ref.) | 6/112 (5.4) | 1.00 | | 8/112 (7.1) | 1.00 | | 9/112 (8.0) | 1.00 | |
| University or above | 76/311 (24.4) | 5.71 (2.41–13.53) | <0.001 | 66/311 (21.2) | 3.50 (1.62–7.55) | 0.001 | 107/311 (34.4) | 6.00 (2.92–12.34) | <0.001 |
| Professional experience (per year) | | 1.06 (1.02–1.11) | 0.015 | | 1.04 (1.00–1.08) | 0.006 | | 1.03 (0.99–1.07) | 0.100 |
| **Professional qualification** | | | | | | | | | |
| Non-pharmaceutical (ref.) | 49/313 (15.7) | 1.00 | | 53/313 (16.9) | 1.00 | | 77/313 (24.6) | 1.00 | |
| Pharmaceutical | 33/110 (30.0) | 2.31 (1.39–3.84) | 0.001 | 21/110 (19.1) | 1.16 (0.66–2.03) | 0.608 | 39/110 (35.5) | 1.68 (1.05–2.69) | 0.028 |
| **Prior training on antibiotic use and AMR** | | | | | | | | | |
| No (ref.) | 19/142 (13.4) | 1.00 | | 23/142 (16.2) | 1.00 | | 50/142 (35.2) | 1.00 | |
| Yes | 63/281 (22.4) | 1.87 (1.07–3.27) | 0.026 | 51/281 (18.1) | 1.15 (0.67–1.97) | 0.618 | 66/281 (23.5) | 0.56 (0.36–0.88) | 0.011 |
| **Health zone** | | | | | | | | | |
| Bagira (ref.) | 16/95 (16.8) | 1.00 | 0.334 | 10/95 (10.5) | 1.00 | 0.075 | 29/95 (30.5) | 1.00 | 0.089 |
| Ibanda | 34/146 (23.3) | 1.50 (0.77–2.90) | | 32/146 (21.9) | 2.39 (1.11–5.12) | | 47/146 (32.2) | 1.08 (0.62–1.89) | |
| Kadutu | 32/182 (17.6) | 1.05 (0.55–2.04) | | 32/182 (17.6) | 1.81 (0.85–3.87) | | 40/182 (22.0) | 0.64 (0.37–1.12) | |
| **Knowledge level** | | | | | | | | | |
| Poor (ref.) | – | – | – | 39/341 (11.4) | 1.00 | | 79/341 (23.2) | 1.00 | |
| Good | – | – | – | 35/82 (42.7) | 5.77 (3.33–10.00) | <0.001 | 37/82 (45.1) | 2.73 (1.65–4.51) | <0.001 |

| Variable | Good knowledge n/N (%) | Crude OR (95% CI) | *P* | Favourable attitude n/N (%) | Crude OR (95% CI) | *P* | Appropriate practice n/N (%) | Crude OR (95% CI) | *P* |
|---|---|---|---|---|---|---|---|---|---|
| **Attitude level** | | | | | | | | | |
| Unfavourable (ref.) | – | – | – | – | – | – | 67/349 (19.2) | 1.00 | |
| Favourable | – | – | – | – | – | – | 49/74 (66.2) | 8.25 (4.76–14.30) | <0.001 |
| Events / N | 82/423 | | | 74/423 | | | 116/423 | | |

**Table B.** Sensitivity analysis: multivariable models of Table 3 with health zone added (n = 417). Adding health zone did not materially change the estimates for the other variables; workers in Ibanda and Kadutu had higher odds of favourable attitudes than those in Bagira.

| Variable | Good knowledge aOR (95% CI) | Favourable attitude aOR (95% CI) | Appropriate practice aOR (95% CI) |
|---|---|---|---|
| Age (per year) | 1.02 (0.98–1.07) | 1.08 (1.03–1.12) | 1.07 (1.03–1.11) |
| Male sex | 1.54 (0.90–2.63) | 1.34 (0.75–2.38) | 0.66 (0.38–1.16) |
| University or above | 4.35 (1.77–10.70) | 2.10 (0.91–4.84) | 5.58 (2.48–12.58) |
| Experience (per year) | 1.02 (0.97–1.08) | 0.94 (0.88–1.01) | 0.96 (0.90–1.03) |
| Pharmaceutical qualification | 2.35 (1.34–4.10) | 0.99 (0.51–1.90) | 2.08 (1.16–3.73) |
| Prior training | 1.22 (0.67–2.22) | 0.68 (0.36–1.26) | 0.32 (0.18–0.55) |
| Good knowledge | – | 4.87 (2.66–8.92) | 1.23 (0.64–2.36) |
| Favourable attitude | – | – | 7.03 (3.67–13.46) |
| Health zone: Ibanda vs Bagira | 1.49 (0.73–3.04) | 2.75 (1.17–6.44) | 0.86 (0.44–1.68) |
| Health zone: Kadutu vs Bagira | 1.22 (0.59–2.51) | 2.87 (1.22–6.77) | 0.64 (0.32–1.25) |

OR, odds ratio; aOR, adjusted odds ratio; CI, confidence interval; ref., reference category; "–" not applicable.
